# Validity of intraoperative imageless navigation (Naviswiss^™^) for component positioning accuracy in primary total hip arthroplasty: Protocol for a prospective observational cohort study in a single-surgeon practice

**DOI:** 10.1101/2020.01.15.20017756

**Authors:** Nalan Ektas, Corey Scholes, Alejandro M Ruiz, John Ireland

## Abstract

**Introduction:** Optimal outcomes in total hip arthroplasty are dependent on appropriate placement of femoral and acetabular components, with technological advances providing a platform for guiding component placement to reduce the risk of malpositioned components during surgery. This study will validate the intraoperative data captured using a handheld imageless THA navigation system against postoperative measurements of acetabular inclination, anteversion, leg length, and femoral offset on CT radiographs.

**Methods and analysis:** This is a prospective observational cohort study conducted within a single-centre, single-surgeon private practice. Data will be collected for 35 consecutive patients (>18years) undergoing elective THA surgery, from the research registry established at the surgeon’s practice. The primary outcome is the agreement between intraoperative component positioning data captured by the navigation system compared to postoperative measurements using computed tomography (CT). A total of ten CT scans will be re-assessed for inter- and intra-observer reliability. The influence of patient and surgical factors on the accuracy of component position will also be examined with multivariable linear regression.

**Ethics and dissemination:** Ethics approval for this study was provided through a certified ethics committee (Bellberry HREC approval number 2017-07-499). The results of this study will be disseminated through peer-reviewed journals and conference presentations.

**Strengths and limitations of this study:**

- This study will assess the accuracy of an imageless THA navigation system for measurement of component positioning against postoperative computed tomography analysis as gold standard.
- A sample size of n=35 participants will ensure adequate power to detect differences between intraoperative navigation results and postoperative CT measurements.
- A sample size of n=10 will provide adequate confidence to establish intra and inter-observer reliability of postoperative measurements of component positioning via CT analysis.
- This study will enable a mechanism to detect potential discrepancies between the component positioning measurement methods intrinsic to the Naviswiss^™^ device and postoperative CT analysis method and identify any corrective factors required for direct comparison of the two methods.

## INTRODUCTION

Optimal outcomes in total hip arthroplasty (THA) are dependent on appropriate placement of components during surgery [1]. Inappropriate component positioning is associated with loosening and impingement, which contribute to suboptimal gait mechanics and unsatisfactory outcomes reported by patients, as well as an increased risk for the eventual need for revision [2–4]. While several computer or robotic-assisted systems improve positioning compared with traditional freehand techniques [5], the financial investment and time required for adopting such systems have precluded their widespread use. Hand-held, image-free navigation devices such as the Naviswiss™ have been developed as a cost-effective, user-friendly and minimally intrusive alternative, however, there is limited clinical evidence regarding the performance of the system. It is thus necessary to demonstrate that any new system meets appropriate standards of accuracy compared to gold-standard measurements, such as computed tomography. The present study was therefore designed, in the first instance, to confirm the validity of the Naviswiss^™^ handheld image-free navigation device for accurate measurement of THA component positioning intraoperatively in comparison with three-dimensional reconstruction of computed tomography as gold standard.

### Objectives

The primary objective of this study is to determine, in patients undergoing total hip arthroplasty using an anterolateral approach, the concurrent agreement between an imageless navigation system applied intraoperatively and postoperative three-dimensional CT reconstruction for cup inclination, cup version, femoral offset and leg length. The secondary objectives are to assess the relationship between patient factors (age, gender, BMI) and concurrent agreement between imageless navigation and CT analysis for component positioning.

## METHODS AND ANALYSIS

### Design and setting of the study

This is a registry-embedded, observational cohort study conducted within a single-surgeon private practice (Sydney, Australia). Repeated measures of component positioning will be extracted from the device log of image-free navigation used intraoperatively and from computed tomography (CT) scans retrieved from preoperative planning and at the routine 6 week follow-up. A reliability assessment for postoperative measurements of component position from CT measurements will be included in the study design (Figure 1).

**Figure 1:**
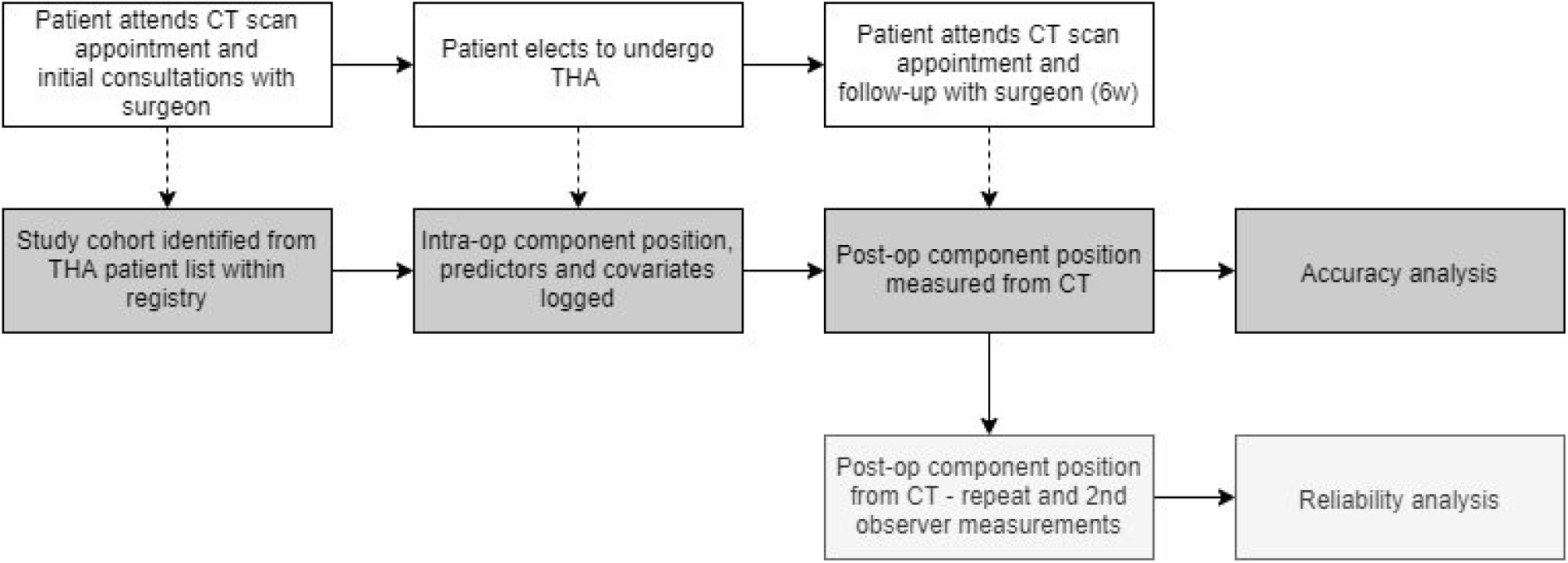
The study design for prospective observational cohort study.

The research is scheduled to commence in January 2020 and due to be completed by December 2020. The study will be registered on an online registry for clinical trials (Australia New Zealand Clinical Trial Registry, ANZCTR), where study site and sponsor details will be listed. The study will be reported as per the STROBE guidelines [6].

### Data sources

The primary data source for collection of identifiers, demographic factors and intraoperative surgical details will be the clinical research registry for hip and knee arthroplasty surgery established within the surgeon’s practice (A prospective assessment of patient outcomes following joint replacement surgery; PAPOJRS; ACTRN12618000317291). Ethical approval for use of the practice registry for research was provided by a National Health and Medical Research Council (NHMRC) certified HREC (Bellberry Ltd). Registry data is hosted in orthopaedic outcome software, located on-site to the clinic (Socrates, v3.5.8.8.10130, MSSQL 2008 R2, Ortholink Pty Ltd, Aus).

Position of acetabular and femoral components intraoperatively will be captured from the case files logged on the Naviswiss^™^ device memory. Postoperative component position will be measured using three-dimensional THA planning software (ZedHip, LEXI Co., Ltd., Tokyo, Japan) and reported in portable document file (PDF) format. Log files and post-operative CT reports will be uploaded to a secure HIPAA-compliant cloud folder. Data from intraoperative Naviswiss^™^ log files and post-op CT reports will be extracted and merged using patient and surgery identifiers to establish a study database that will be used to answer the research questions. Data collection will continue until a total of 35 cases with complete records have been extracted.

### Participants

#### Eligibility and Recruitment

Potential patients will be identified consecutively from the clinical research registry (Figure 2). Adult patients (>18y) are included in the THA cohort of the research registry if they present to the primary investigator with end-stage osteoarthritis or rheumatoid arthritis and elect to undergo THA surgery. Patients who have declined or revoked consent for use of clinical data for research, or unable to provide informed consent are excluded from the registry.

**Figure 2:**
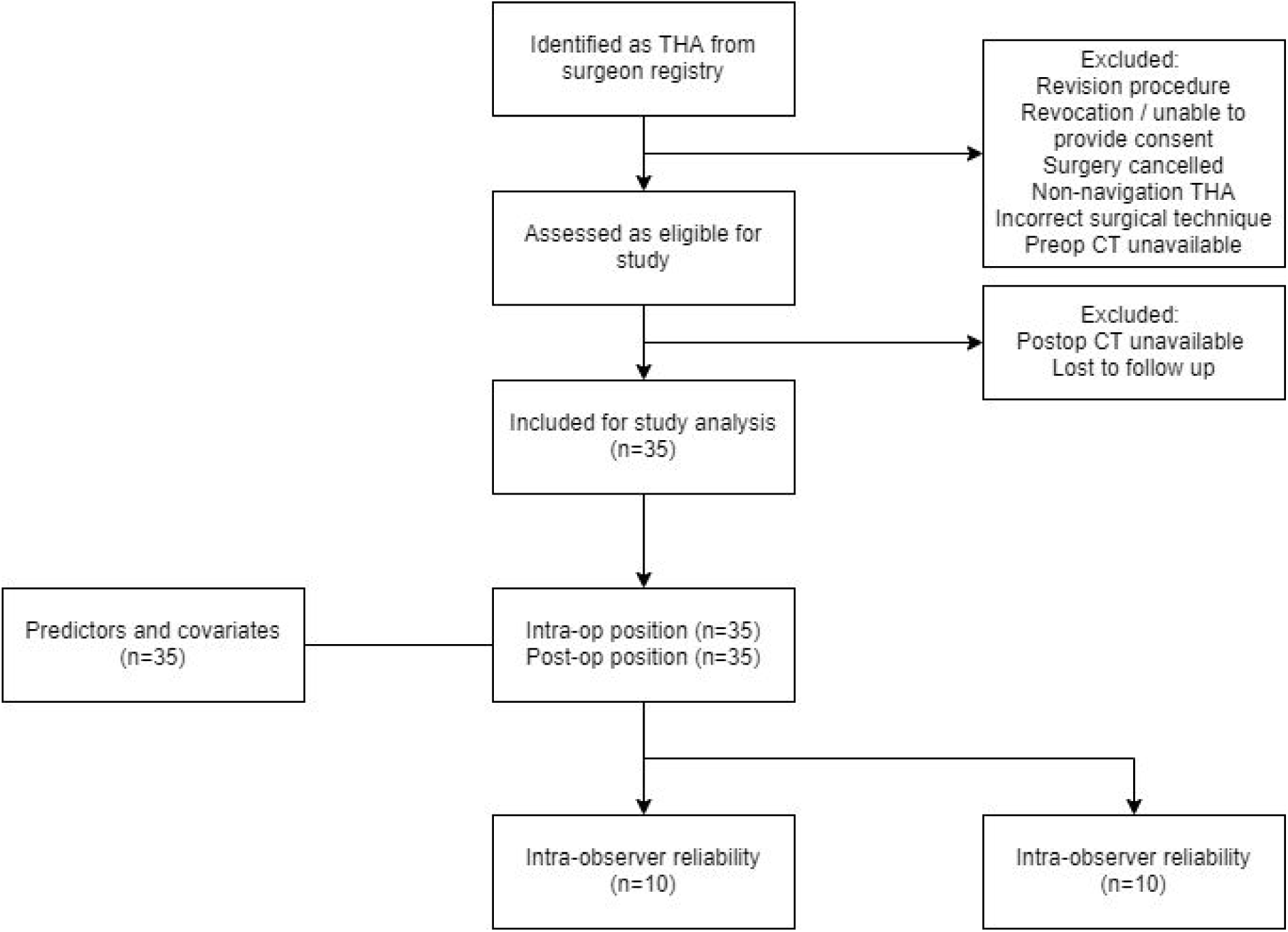
STROBE diagram [6] with key steps that will lead to patients being included in the analysis.

The study cohort will comprise primary THA cases where an anterolateral surgical approach and supine position was used, and for whom postoperative CT scans are available via the picture archiving and communication system (PACS) following the routine 6 week follow up. Patients are referred to either of two imaging providers depending on the location of their surgery. Cases in which a short-stem component was implanted, ipsilateral revision procedures, simultaneous bilateral procedures or cases involving severe contralateral hip deformity or dysplasia will be excluded from this study.

The use of the imageless navigation system is currently standard practice for the participating surgeon and consent is established from all participants for contribution of de-identified clinical data to the clinical research registry via an opt-out approach [7]. Eligible patients will be presented with a Patient Information Sheet and a Withdrawal of Consent form by the consultant surgeon or practice nurse. Patients are given the opportunity to ask any questions, and opt-out of research if they so wish by completing the Withdrawal of Consent form.

#### Power Analysis

The sample size was established to provide adequate power to detect a 2.5° absolute mean difference between intraoperative navigation results and postoperative CT measurements for inclination and version using a paired t-test design (each patient acts as their own control), with an assumed between-patient standard deviation of 5°. The standard deviation was estimated from an initial pilot study with 15 cases during the learning curve (2.4° for inclination and 2° for version) with an additional margin of error added. Power (□) was selected at 0.8 and α of 0.05 with an estimated sample size of 34 cases.

The sample size necessary for the multivariable regression analysis (N = 30) with patient *age at surgery, sex* and *body mass index* selected as the model predictors was estimated from a model R^2^ of 0.3, three predictors and the same □ and α.

The sample size necessary to establish intra and inter-observer reliability of the post-operative CT measurements (N = 10) was estimated from a two-sided test of correlation within and between observers, with an R^2^ of 0.5 and a beta/alpha ratio of 1. All power calculations were performed with a dedicated software package (Gpower v3.1.9.2, University of Kiel, Germany). A sample of 10 cases also provides a 95% confidence interval around the primary and secondary observer average estimates with a 95% confidence interval around a margin of error of 3° (version/inclination) and a standard deviation of 5°, using the formula;

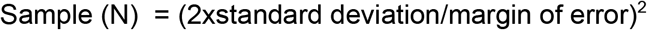

### Outcome measures

#### Primary study outcomes

The following variables will be extracted from intraoperative data captured on the Naviswiss^™^ device and measured postoperatively on CT scans to identify primary study outcomes:

- Acetabular cup inclination (ACI),
- Acetabular cup anteversion (ACV),
- Femoral offset (FO), and
- Leg length (LL).

The following demographic factors or intraoperative/surgical details will be collected for data linkage and to identify factors that potentially affect the accuracy of component positioning:

- Patient demographics
  - Age at surgery, gender, height, weight

- Potential factors associated with component positioning accuracy
  - Patient factors: BMI, diagnosis
  - Surgical factors: surgical approach, extraction table, hardware/software versions, case declarations

- Identifiers for data linkage and error tracking
  - Patient ID, surgery date, surgery start and end times, surgery duration, data collection rep, case and log files, imaging details

### Instrumentation

The Naviswiss^™^ system (Naviswiss AG, Switzerland) is an image-free surgical navigation system, which assists the orthopaedic surgeon during the THA procedures. It consists of a handheld navigation device that is used to register the patient’s anatomy. Subsequently the navigation system supports the surgeon to guide the surgical instruments with the goal to position the implant according to the pre-operative plan. The navigation unit includes an infrared stereo camera that measures the position and orientation of small tags mounted to the pelvis and the greater trochanter of the femur. The tags are mounted with bone pins to provide pelvic and femoral orientation data to the camera unit. An infrared flash illuminates the field of view to avoid available-light disturbance. An inertial measurement unit (IMU) is built-in which measures the camera orientation in space. A caliper is used to identify the anterior superior iliac spine bilaterally to establish the functional pelvic plane, combined with IMU data to establish the gravitational axis and embed a coordinate system into the pelvis during the procedure. The system is registered as a Class IIa device on the Australian Register of Therapeutic Goods (ARTG305085), and thus conforms to all Essential Principles (including safety) and is subject to continued post-market monitoring.

### Surgical technique and intraoperative measurement of component positioning

All surgeries will be performed by the senior author. Preoperative templating will be performed using 3D CT assessment and functional views for estimation of appropriate femoral and acetabular component sizing and orientation [8]. The surgery will be performed with the patient in supine position with exposure to the hip via an anterolateral approach using a specialised extraction table designed to facilitate femoral exposure. An incision will be made posterior and distal from the anterior iliac spine, extending distally over the belly of the tensor fascia latae [9]. Guidance for component positioning will be provided intraoperatively using the image-free navigation device. With sterile, single-use tags fixed to the pelvis and greater trochanter using intracortical bone pins, the caliper set is applied to the ASIS bilaterally to establish the functional pelvic plane (FPP). The hip centre of rotation is identified by the functional method [10], with the thigh moved through a multiplanar range of motion while the tags are tracked with the handheld camera. The femoral head will be resected for femoral and acetabular exposure. The acetabulum will be reamed and the acetabular component (Novation Crown Cup, Exactech, USA) placed as per manufacturer’s instructions and the orientation confirmed with the navigation device. The femoral canal will be prepared as per manufacturer’s instructions. The femur tag will be fitted, the hip reduced and positioning of the femur checked using the navigation device with the leg in neutral position. Adjustments to stem or head sizing will be made using the navigation system for guidance, followed by insertion and impaction of the appropriate sized components. The final intraoperative component positions (cup inclination, cup anteversion, leg length and femoral offset) will be checked in neutral position, logged by the navigation system and exported for analysis.

### Measurement of component positioning

Blinded DICOMs will be used for all postoperative CT measurements of component positioning, with information relating to the specific diagnosis, study, surgeon or whether navigation was used for the hip arthroplasty procedure removed prior to measurement of component position. The postoperative images will be blinded by an independent research assistant who will not be involved in performing the measurements.

Postoperative component position will be measured by uploading DICOM data to dedicated software (ZedHip, Lexi Co Ltd, Japan and Imarti, Imatri Medical, South Africa) to measure anteversion and inclination of the acetabular cup, FO and LL through assessment of the geometric characteristics of the hip and THA components using 2D/3D visualisation and MPR (Multi-Planar Reconstruction) functions. For the postoperative CT assessments, coordinate systems for the pelvis and femur will be determined using the anatomic and ISB coordinate systems respectively. Parameters from both the Naviswiss^™^ and CT analysis system will be expressed relative to the functional pelvic plane, with the origin placed at the centre of the line connecting the left and right anterior superior iliac spine (ASIS). Parameters for acetabular orientation will be determined as previously described [11], with inclination defined as the angle between the acetabular and longitudinal axes when projected onto the coronal plane. Cup anteversion will be measured as the angle between the acetabular axis and coronal plane. Femoral offset is defined in the navigation system as the relative difference between the hip centre of rotation (COR) of the operated joint relative to its starting position at the initial assessment in the coronal plane (medial-lateral) within the pelvic coordinate system. A similar definition is applied for leg length, with the change in the distance between the greater trochanter tag and the hip COR summed with the change in the distance between the centre of the acetabulum and the centre of the cup in the transverse plane (superior-inferior) reported by the navigation system. For the postoperative CT analysis, the position of the cup centre will be compared to the native hip centre of rotation (COR) determined from the preoperative CT. Femoral offset and leg length will be reported as the pre-to-post change in COR coordinates in the coronal (medio-lateral) and transverse (inferior-superior) planes respectively.

### Statistical Analysis

Data will be exported from the clinical research database and cross-matched to the practice management system to identify consecutive eligible patients for the study cohort. The characteristics of study cohort will be summarised in a STROBE (Strengthening the Reporting of Observational studies in Epidemiology) diagram [6] which will illustrate the eligibility and inclusion of patients captured for the study and report rates of data completeness. Differences between intraoperative and postoperative measurements of the primary outcome variables (ACI, ACA, FO and LL) will be used to determine the limits of agreement, and validate the accuracy of the Naviswiss^™^ system. Reliability of postoperative measurement of component positioning will be conducted via inter and intra-rater reliability assessments. Intra-rater reliability will be established with a subset of cases (N = 10) randomly selected with a random number generation (Matlab 2018b, Mathworks Inc, USA) for repeat measurement of component positioning by the primary observer. Variability in the primary observer’s measurements will be assessed by calculating the standard error of measurement and Bland-Altman limits of agreement [12]. Repeat measurements will be conducted at a minimum of 2 weeks or more after the first measurement [13], with the primary observer blinded from the initial measurement values. Inter-rater reliability will be determined by a second independent observer measuring the same repeated cases. The second observer will be blinded from the primary observers measurements. The reliability of observer measurements will be assessed with intra-class correlation and standard error of measurement [14].

The intraoperative data and postoperative CT measurements of the primary outcomes by the primary observer will be compared using mixed effects linear regression [12]. To account for the technical error associated with CT, the reliability measurements derived from the subset selected for repeat assessment will be used to simulate repeat measurement data for the remaining cases (N = 25), assuming a normal distribution of variation around the average difference between measurements for the same observer. A random assignment between the simulated dataset and the remaining cases will be conducted using a random number generator. The model results will be then estimate the differences between methods while accounting for technical error in the partitioning of variance, with measurement method as a fixed effect and patient identifier as a random effect [15]. Residual analysis from the model will be used to determine the standard error of measurement of the navigation system relative to CT measurements (Hopkins), as well as the proportion of cases where navigation and CT measurements are within 3° of each other or 5mm for FO and LL [14]. The secondary analysis will be addressed within a second mixed effects model of the residuals extracted from the first model and including gender as a fixed effect, with age at surgery and body mass index included as covariates to establish their contribution to the differences between methods. The statistical analyses will be performed in dedicated software (Minitab v18, Minitab Inc, USA) with α of 5%. Partial *η*^2^ will be calculated to report the size of effects for model factors.

### Study Termination

To ensure that patients are not exposed to inaccurate guidance, accuracy criteria have been established by the investigators prior to study commencement. An online dashboard displaying aggregated de-identified data has been established for the investigator team and sponsor representative to monitor whether the difference between the intraoperative data and the primary observer’s first postoperative measurement exceeds 10° for cup inclination and version and 10mm for femoral offset and leg length. The study will be paused if greater than 5% of cases (2 out of 35) exceed these limits for each variable, without a declaration logged during intraoperative data collection. A case is considered *declared* if an issue with the measurement process is observed in-theatre and logged on a pre-specified electronic form. The cases exceeding the threshold will be investigated and the study terminated if the discrepancies cannot be attributed. The thresholds were established from one study that reported 97% of cases within 10° between an imageless navigation system (Orthoalign^™^) and CT measurements in 75 cases operated in the supine position, including the learning curve [16]. A second study reported 100% of cases (N = 25) within 10mm for leg length between navigation (Intellijoint^™^) and radiographic measurements [17].

### Ethics And Dissemination

The proposed trial is embedded within a prospective observational practice registry (ACTRN12618000317291) with HREC approval (Bellberry Limited 2017-07-499). Findings will be disseminated through peer-reviewed journals and national and international conferences using aggregated de-identified data so as to protect the privacy and confidentiality of participants. Authorship eligibility will be determined as per ICMJE guidelines, and all sources of input for the final publications will be acknowledged.

### Availability of data and materials

The datasets generated and/or analysed during the current study will not be publicly available due to reasons of confidentiality and commercial-in-confidence. Deidentified data may be made available from the corresponding author on reasonable request.

### Documenting protocol amendments

Amendments to the study protocol will be documented within the final study manuscript. The nature of the changes will be agreed to by study investigators and stakeholders. Relevant sections of the study registration record will be updated on the Australia New Zealand Clinical Trial Registry as appropriate.

## DISCUSSION

This study will provide, in the first instance, clinical data pertaining to the validity of a newly introduced imageless navigation system (Naviswiss AG) for determining accurate component positioning in THA through comparison with three-dimensional reconstruction of computed tomography as a criterion gold standard. CT based measurements of postoperative component position following THA have been compared to 2D radiological measurement of cup position [18,19] imageless [20,21] or accelerometer-based navigation THA systems [22]. However there is limited information on the reliability of CT-based measurements in the context of its validation as a gold standard. While one study assessed the precision (defined as the level of agreement between repeated measurements) and bias (consistency between sets of measurements) of navigation systems with respect to CT evaluation [21], the reliability of the CT measurements themselves were not reported. The intra and inter-observer assessments adopted within this protocol will assist in validating the navigation system in the context of CT measurement error.

This study will also examine factors which may influence the accurate placement of components intraoperatively, and may potentially identify a subset of patients or surgical factors that increases the probability of favourable component positioning, and inform selection of patients suitable for use with the imageless guidance system. With patient anthropometry, in particular BMI and hip anatomy, a known factor predicting outcomes following THA [23], this study will confirm whether the imageless navigation system can achieve successful component placement through identification of superficial anatomical landmarks in patients with high BMI or complex hip anatomy.

With the goal of THA to restore the normal biomechanics of the hip joint [24], correct identification of the hip joint centre, is required for measurement of the femoral offset and leg length [25]. However, inconsistencies in the reporting of coordinate systems for joint mechanics have been previously identified, with the lack of standard reporting resulting in difficulties with direct comparison amongst studies or measurement methods [26] Previous validation studies have investigated a variety of methodologies, and identified differences in measurements of hip joint centre coordinates of 13mm, and up to 25-30mm on average for functional or prediction methods respectively, compared to roentgen stereophotogrammetric analysis (RSA). [10] While the functional approach for determining the hip joint centre is recommended [26]), examination of motion of the hip is required, which may exclude some patients, or assessments where physical evaluation is unavailable, such as with postoperative CT analysis of hip component position. The authors note that while attempts have been made to replicate the measurement methods for the navigation system within the CT analysis, inherent differences in the calculation of variables may be unavoidable. This study will enable a mechanism to detect potential discrepancies between the measurement methods intrinsic to the Naviswiss^™^ device, whereby the hip centre of rotation is determined using the functional method, and the CT analysis methods, and identify any corrective factors required for direct comparison of the two methods proposed.

## Data Availability

As this is a study protocol, there is no data available.

## Authors’ contributions

NE, CS and JI have contributed to the conception and design of the study. NE and CS contributed to drafting the protocol, revisions of the manuscript and will draft the study report. All authors were involved in critically revising and approving the final version of the manuscript.

## Version History

v1.0 (16/1/2020): Original protocol finalised

v1.1 (10/3/2020): Minor adjustment to Methods section; addition of new software for 2D/3D analysis.

